# On the hip abduction moment required to balance on one leg: an experimental study

**DOI:** 10.1101/2022.04.14.22273888

**Authors:** Payam Mirshams Shahshahani, Mariana Masteling, James A. Ashton-Miller

## Abstract

Although the time a patient can stand on one leg is a common clinical test of balance in those prone to fall, a surprising knowledge gap is how much hip abduction muscle strength is required. This is important because hip abduction strength has been shown to be important for compensating for impairments in diabetic neuropathy, for example. As a start we tested the hypothesis that maximum hip abduction muscle endurance time at 50% effort would be longer than the time that 18 young and 17 older healthy adults can stand on one leg. First, maximum hip abduction endurance time at 50% effort as well as maximum abduction strength were measured in the gravity-free plane. Then subjects were asked to balance on their left foot for as long as they could while body segment kinematics and ground reaction data were measured. The results showed that the mean intensity of the hip abduction moment required to stand on one leg exceeded 50% of the maximum hip abduction strength for all four groups (young women and men 53% and 55%, and older women and men 94% and 72% respectively). However, unipedal stance times were not limited by hip abduction 50% effort endurance time (p = 0.9). Therefore a significant portion of the hip abduction moment required to stand on one leg must be carried by passive tissues. The underlying mechanism remains to be explained.

## 2. Introduction

One-legged balance (OLB) is one of the most commonly used clinical tests of balance in the elderly [1,2]. The clinician asks the patient to stand on one leg for as long as possible and measures the maximum unipedal stance time (UST) as the primary outcome measure [3,4]. The patient passes the test if the UST exceeds 5s [3] or, alternatively, 30s [5]. Patients who fail the OLB test are considered at greater risk of future injurious falls [3,5]. In previous studies, we showed that, in a cohort of patients with diabetic peripheral neuropathy, their maximum hip abduction strength could compensate for the loss of ankle sensorimotor function in determining both the UST as well as the risk of future injurious falls [6,7]. Hip abduction strength is therefore important. But a current knowledge gap is that the hip abduction strength requirement for OLB is unknown, which means one does not know how much to strengthen the abductor muscles in order to improve UST. This is partly because the focus of most biomechanical studies of OLB has been distally on the sensorimotor function of the stance ankle and foot, rather than proximally on the demands on the ipsilateral (stance) hip muscles [8,9].

In an earlier paper we used a simple double inverted pendulum mass-link model to estimate the ankle and hip strength frontal plane requirements for OLB [10]. One link represented the stance leg while the other represented the rest of the body. For simplicity, we assumed that the rest of the body was kept rigid and the stance foot kept stationary. Starting with the anthropometric parameters of a mid-size male [11] and scaled published ankle inversion and eversion, as well as hip abduction strengths, from the literature [7,12,13], we calculated that the hip abduction moment demand required for quasistatic OLB exceeds 50% of the maximum voluntary hip abduction strength for all adult groups (healthy young, healthy older, as well as older patients with diabetic peripheral neuropathy).

Based on the above result as well as the known fatigue behavior of striated muscle during a sustained isometric contraction [14], we hypothesized that endurance of the hip abductor muscles (mostly the gluteus medius, gluteus minimus, and tensor fasciae latae) may limit maximum unipedal stance time. A limitation of this argument is that our estimation of the intensity of the hip abduction moment demand for OLB had to be based on non-coherent published strength and anthropometric data from a variety of sources. So in this paper we address this limitation directly by conducting a timed OLB experiment in young and older adults while also gathering subject-specific estimates of the intensity of hip abduction moment demand.

We tested the hypothesis that measured endurance time at a 50% of maximum hip abduction strength exertion exceeds the measured UST. Alternatively, the calculated endurance time based on subject-specific hip abduction moment demand intensity will reliably exceed the maximum UST for that subject. This hypothesis relies on the assumption that, in healthy adults, the hip abduction moment required for OLB is completely developed by the active abductor muscles. Rejection of these hypotheses would imply that there is a significant hip abductor moment contribution from other sources in OLB.

## 3. Methods

### 3.1. Participants

Each participant gave written informed consent (Institutional approval # HUM00130970). Recruitment details can be found in S1 Text Section 1. The demographics of the participant pool and their unipedal stance times (UST) are provided in Table 1.

**Table 1.** Mean (standard deviation) demographics and unipedal stance time (UST) in the OLB experiments. The p-values are from two-way ANOVA tests with age group and sex as independent variables. UST distribution was highly non-normal due to censoring because many participants reached the maximum trial time of 4 minutes. Therefore, two-way ANOVA was not performed for UST.

|  | *Young (20-30 years)* |  | *Older (65-80 years)* |  | *p-value* |
| --- | --- | --- | --- | --- | --- |
|  | *Men*<br>*(n = 8)* | *Women*<br>*(n = 10)* | *Men*<br>*(n = 7)* | *Women*<br>*(n = 10)* |  |
| *Age (years)* | 25 (2.5) | 26 (2.8) | 70 (4.0) | 69 (3.6) | < 0.001<sup>1</sup> |
| *Weight (Kg)* | 76.1 (11.4) | 62.5 (6.6) | 82.4 (5.3) | 64.6 (8.1) | < 0.001<sup>2</sup> |
| *Height (cm)* | 174 (5) | 168 (6) | 177 (5) | 159 (5) | 0.01<sup>3</sup> |
| *BMI (Kg/m<sup>2</sup>)* | 24.9 (3.1) | 22.2 (1.8) | 26.4 (2.2) | 25.4 (2.9) | 0.007<sup>1</sup><br>0.03<sup>2</sup> |
| *UST (s)* | 167 (89) | 230 (22) | 71 (80) | 57 (37) | -- |

<sup>1</sup>Age main group effect
<sup>2</sup>Sex main group effect
<sup>3</sup>Age:Sex group interaction

### 3.2. Testing Sequence

All the subjects were healthy and declared on a screening survey they had no neuromuscular or musculoskeletal conditions or medications that affected their balance (S1 Text Section 1). They participated in both the OLB experiment and a later volitional heel-toe-shuffle (HTS) experiment in the same testing session (**Fig 1**), but in this paper we shall only report on the OLB experiment.

**Fig 1.**
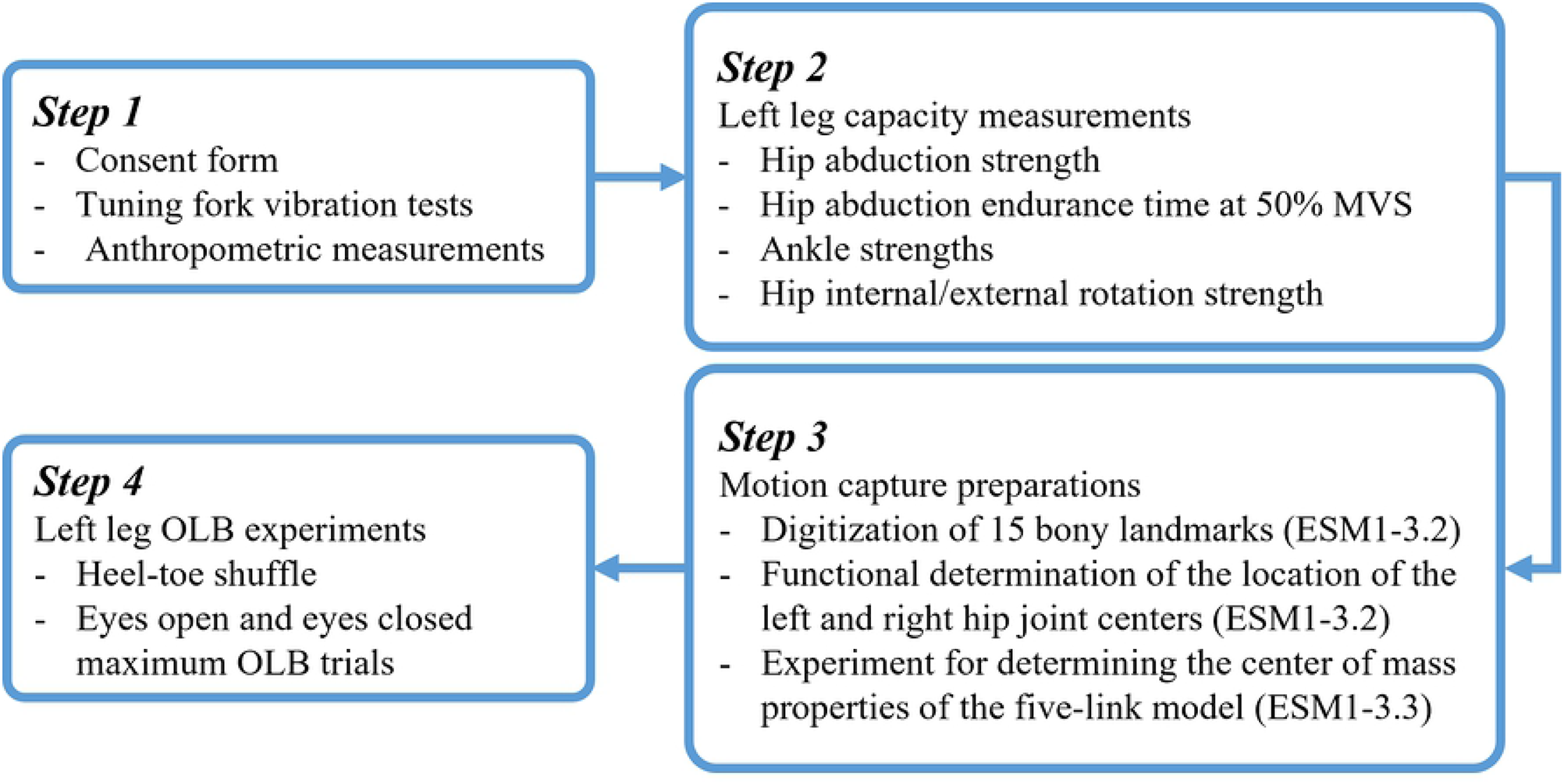
Overview of the sequence of measurements and experiments for each participant.

#### Left leg isometric hip muscle strength measurements

Left hip abduction maximum voluntary strength (MVS) and endurance time at a target of 50% MVS were measured in supine position and gravity-free plane. Ankle strength measurements were performed while the participant stood on their left foot on a 1,000 lbf capacity AMTI force plate (Optima HPS) sampled at 500 Hz. The ankle inversion and eversion strengths were calculated by multiplying the weight of the participant times the maximum volitional center of pressure (COP) deviations from neutral (defined as a line between the second metatarsal joint and heel) in the lateral and medial directions, respectively. The participant was allowed to momentarily touch safety hand rails on either side to recover balance if necessary during the testing. Full details of our protocol for measuring left leg isometric strengths and a table of the measured left leg strengths can be found in S1 Text Section 2.

The ‘adjusted’ maximum hip abduction strength for each participant was then calculated as the larger of two values, either: 1) the measured hip abductor MVS, or 2) the MVS calculated from the estimation of the ‘true intensity’ of the hip abduction moment from the hip abduction endurance time test. The second value was based on published joint-specific endurance time versus intensity curves for young and older adults [14,15]. This adjustment increased the hip abduction strength value for five of younger participants (out of 18) and eight of the older participants (out of 17). We used this adjustment due to the difficulty that some subjects had in coordinating their hip muscles to perform a maximum isometric hip abduction effort in the supine position versus the more familiar task of maintaining enough hip abduction effort to keep a weight sized to act as a 50% MVS of each participant suspended off the ground. Both sets of results will be provided. Details of the calculations and in-depth reasoning behind this adjustment are provided in S1 Text Section 2.

#### Maximum one-legged balance time trials

All OLB tests were performed on the left leg which allowed the lab-based Certus (Northern Digital Inc., Waterloo, ONT, Canada) motion capture setup to reliably track all optoelectronic kinematic markers during the trial at 90 Hz. All but five of the participants wore New Balance 411 walking shoes provided by the Biomechanics Research Lab. The exceptions were for the participants whose shoes size was not available in the lab. Before the testing started, each participant received the same instructions: “try to keep your back straight and arms crossed at all times; your right leg and foot should not touch the left leg at any point”. Movements of the stance foot were allowed as long as the foot stayed entirely within the bounds of the force plate. If the right foot touched the ground, or the participant maximally bent their trunk in the lateral or anterior direction, the test was halted. The examiner either halted data recording at the four minute mark or upon failure to stand on one leg under the conditions described above, whichever was first. Each participant performed OLB trials both with eyes open and with eyes closed. Since OLB with eyes closed involves more dynamic movements than OLB with eyes open, and our hypotheses are concerned with the strength requirements of quasistatic OLB, in this paper, we will only examine the data from the eyes open trials.

### 3.3. Calculating the ankle moment and hip abduction moment and intensity during the OLB experiment

For the calculation of ankle and hip moments during OLB we used a simple double inverted pendulum model to represent the subject in the frontal plane [10]. Full description of how the parameters and states of the double inverted pendulum model were calculated can be found in S1 Text Section 3. The ankle eversion and hip abduction moments at each instant were calculated using a weighted least squares method for the inverse dynamic analysis described by van den Bogert and Su [16].

To calculate the intensity of the OLB hip abduction moment demand, we divided the hip abduction moment demand by the maximum isometric strength at any time point thereby deriving a time-series for the intensity of the hip abduction moment demand (and expressed as a percentage). We accounted for the change in maximum isometric hip abduction strength with the hip abduction angle as described earlier [10].

### 3.4. Statistical Analyses

All statistical analyses were performed with R version 3.6.0 and an alpha level of 0.05 was considered significant. The kinematic and inverse dynamic calculations and linear regressions were performed with MATLAB © version R2018b. Hip abduction moment demand and intensity time-series for each participant were fitted by linear regression with time as the independent variable. The slope of the regression line, root mean square error (RMSE) between the fitted line and the data, as well as trimmed mean without 20% of the outliers were extracted for each participant’s time-series S1 Text Section 3.7. Subsequently, two-way ANOVA was used for between-group comparisons between age group and sex. A two sided paired student t-test was used to test the main hypothesis with p<0.05 being considered significant.

## 4. Results

### 4.1 Estimated hip abduction moment demand and intensity during OLB experiment

The trial in **Fig 2** is representative of all participants who stood for more than 20 seconds. We can see that while there are small variations in both the moment and intensity values, the general trend is linear. Both the intensity of the hip abduction moment demand and RMSE of the data with the trend line were significantly higher in the older group (**Table 2**). This difference is mostly driven by the difference in maximum voluntary hip abduction strength between young and older adults. The slopes of the changes in intensity were not significantly different from zero indicating that even for the long OLB trials, our participants did little to alleviate the high hip abduction moment intensity.

**Fig 2.**
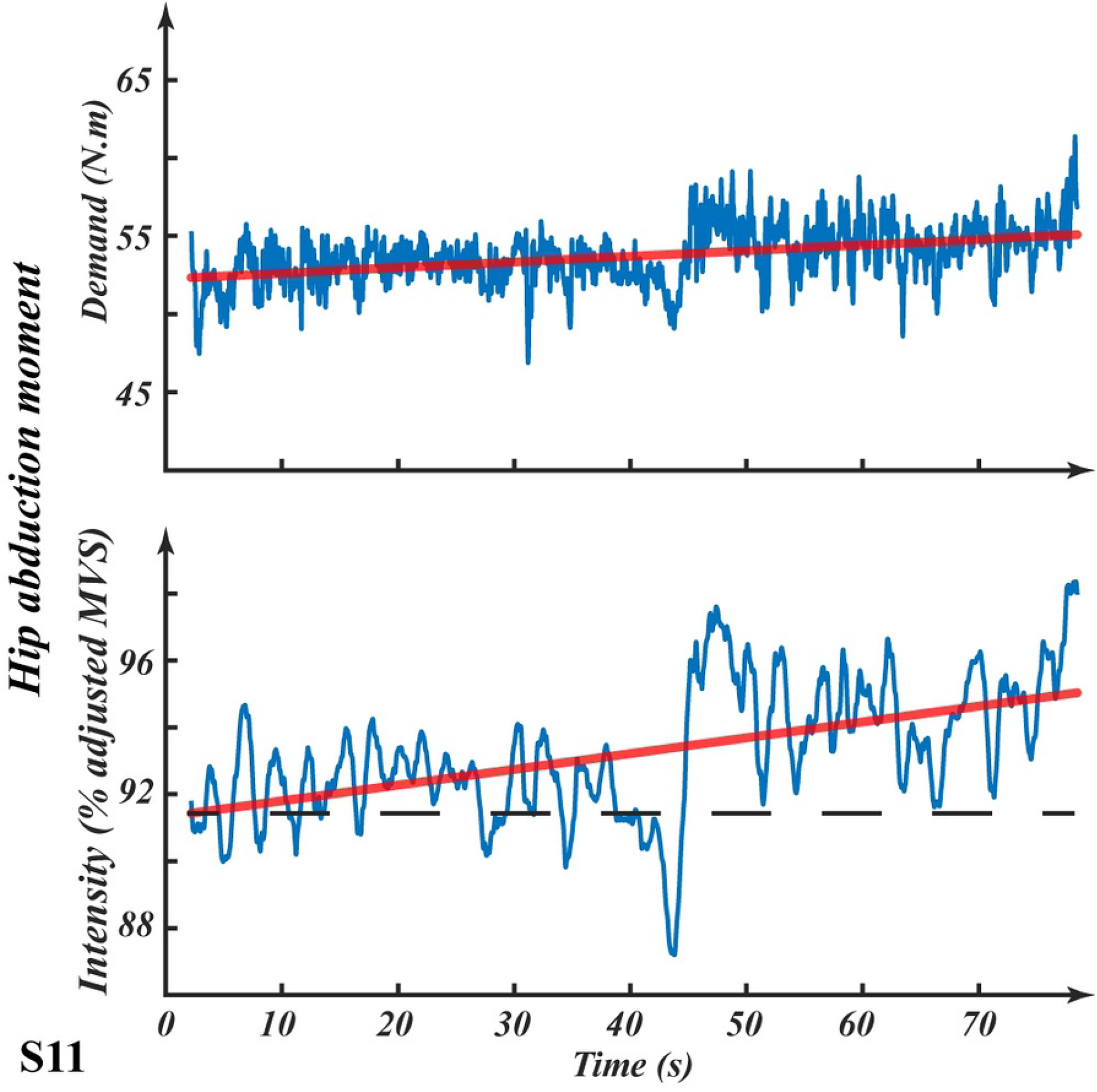
Sample time-series for calculated hip abduction moment (top) and intensity of hip abduction moment demand (bottom) during a OLB trial for subject S11. The solid red line in both plots is the linear regression model fitted to the time-series. Note that neither ordinal axis begins at zero.

**Table 2.**
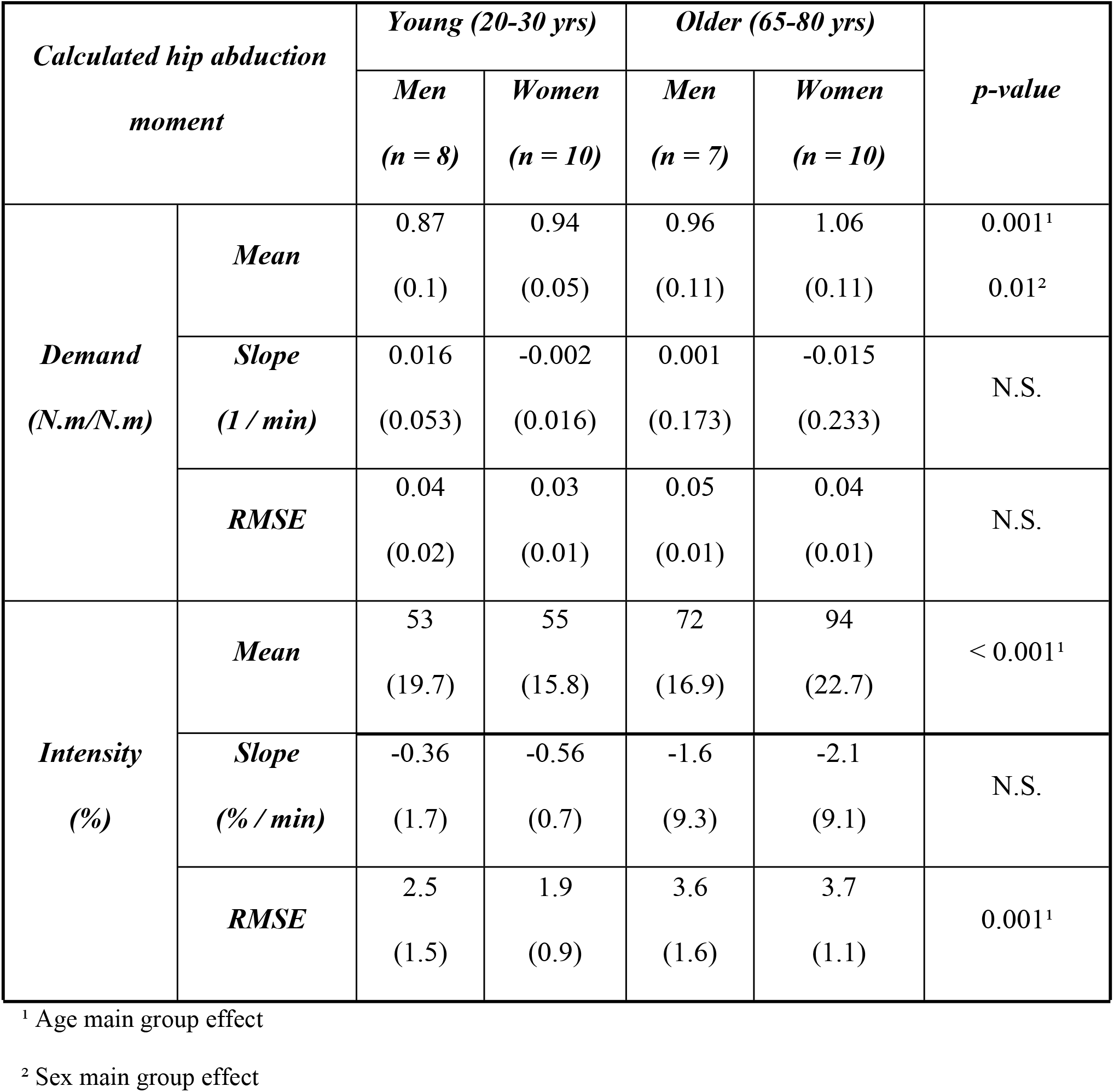
Descriptive statistics for the time-series of calculated hip abduction moment demand (normalized by weight and half the inter-acetabular distance, please see S1 Text Section 3.6), and intensity of the hip abduction moment demand (% of adjusted maximum voluntary hip abduction strength) for all participants.

| **Calculated hip abduction**<br>**moment** |  | **Young (20-30 yrs)** |  | **Older (65-80 yrs)** |  | **p-value** |
| --- | --- | --- | --- | --- | --- | --- |
|  |  | **Men**<br>**(n = 8)** | **Women**<br>**(n = 10)** | **Men**<br>**(n = 7)** | **Women**<br>**(n = 10)** |  |
| **Demand**<br>**(N.m/N.m)** | **Mean** | 0.87<br>(0.1) | 0.94<br>(0.05) | 0.96<br>(0.11) | 1.06<br>(0.11) | 0.001<sup>1</sup><br>0.01<sup>2</sup> |
|  | **Slope**<br>**(1 / min)** | 0.016<br>(0.053) | -0.002<br>(0.016) | 0.001<br>(0.173) | -0.015<br>(0.233) | N.S. |
|  | **RMSE** | 0.04<br>(0.02) | 0.03<br>(0.01) | 0.05<br>(0.01) | 0.04<br>(0.01) | N.S. |
| **Intensity**<br>**(%)** | **Mean** | 53<br>(19.7) | 55<br>(15.8) | 72<br>(16.9) | 94<br>(22.7) | < 0.001<sup>1</sup> |
|  | **Slope**<br>**(% / min)** | -0.36<br>(1.7) | -0.56<br>(0.7) | -1.6<br>(9.3) | -2.1<br>(9.1) | N.S. |
|  | **RMSE** | 2.5<br>(1.5) | 1.9<br>(0.9) | 3.6<br>(1.6) | 3.7<br>(1.1) | 0.001<sup>1</sup> |

<sup>1</sup> Age main group effect
<sup>2</sup> Sex main group effect

### 4.2. Relationship between hip abductor muscle endurance times and UST

All of the young participants and four of the older participants had a longer UST that their own endurance time measured at half of their maximum hip abduction strength (**Fig 3A**). In the test of the main hypothesis (UST is shorter than the measured endurance time for each subject at 50% effort), the paired student t-test revealed that the measured endurance time was not significantly longer than the UST (p = 0.9). This implies that the subjects in our cohort were using less than 50% of their maximum voluntary strength to stand on one leg. Alternatively, the estimated endurance times for each subject based on their calculated effort did not change that result (**Fig 3B)**. In summary, most of the participants stood on one leg for much longer than the predicted hip abduction endurance times.

**Fig 3.**
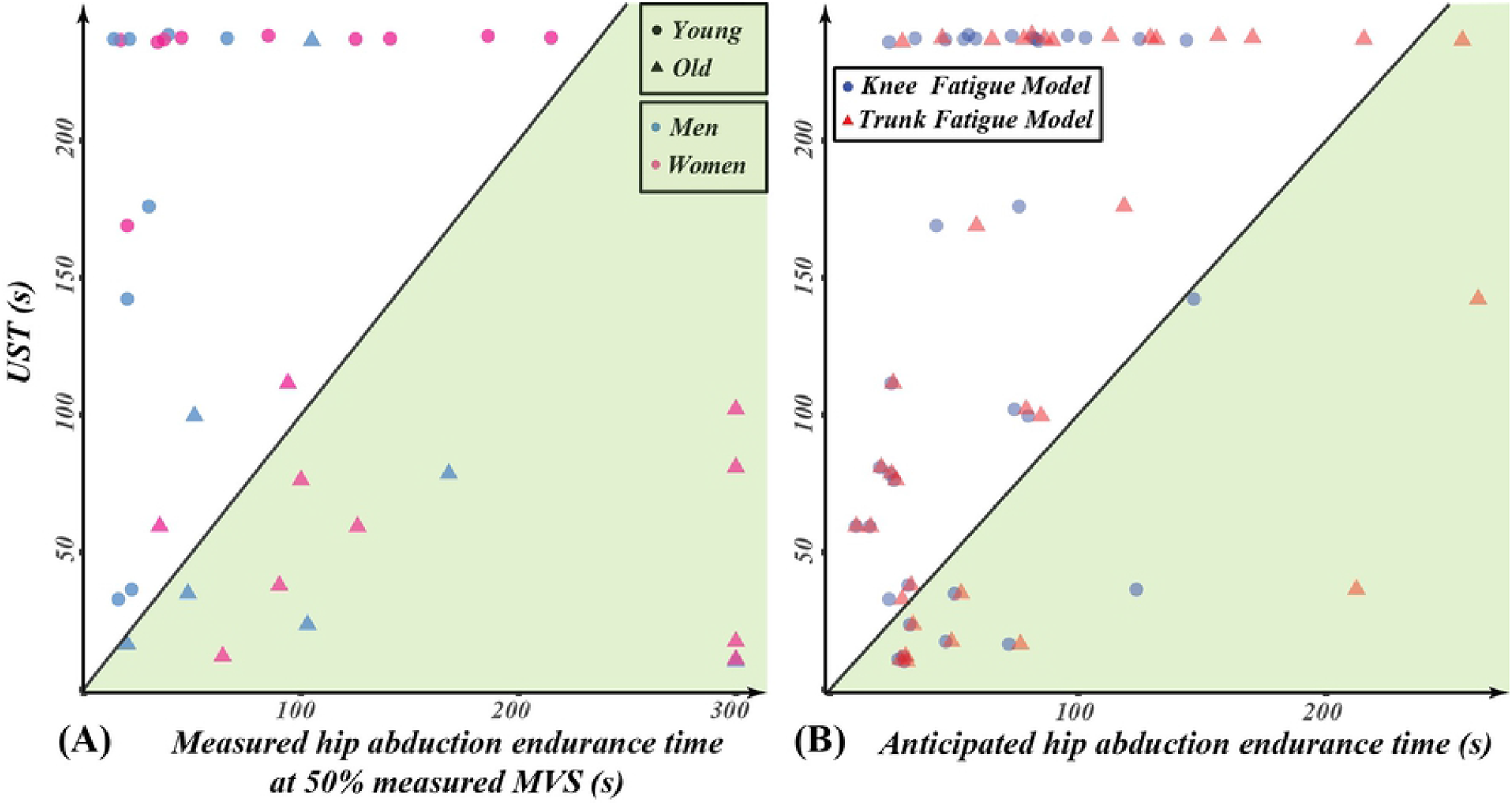
Comparison of measured unipedal stance time (UST, on y-axis) with measured and anticipated endurance times. **(A)** The horizontal axis is the endurance time measured at half of the participant’s measured MVS. If participants used more than 50% of their hip abduction MVS during OLB trial, then one would expect their UST to lie in the green region below the diagonal line having a zero intercept and slope equal to one. **(B)** The horizontal axis is the anticipated max UST based on the calculated mean intensity of the hip abduction moment demand during the OLB trial for each participant and the published isometric exertion endurance time model parameters for knee and trunk [14,15]. Knee and trunk fatigue models were used because there are not enough endurance time studies for the hip abductor muscles in the literature. If all the hip abduction moment demand during OLB was provided by the active hip abductor muscles, then the fatiguing of these muscles would have to maintain the UST in the green region.

## 5. Discussion

To our knowledge this is the first study to quantify experimentally the subject-specific intensity of the hip abduction moment demand required for OLB. In so doing there is no doubt that the hip abduction demand to maintain OLB is strenuous: the calculated abduction intensity exceeded 50% of the adjusted maximum supine hip abduction strength in both the young and older adults (**Table 2**). A strength is that this finding corroborates our earlier estimates [10] based on non-coherent published strength and anthropometric data. A second novel finding was that, despite the large calculated abduction intensity, our results still led to the rejection of the primary hypothesis: that neither the measured endurance time of the hip abductor muscles at half-maximum hip abduction MVS nor the hip abduction endurance time predicted from the intensity of the hip abduction moment demand predicted an upper limit for the UST.

The rejection of our hypothesis was surprising because, like any other human balance task, sufficient joint strength is only one of many factors required to balance on one leg for long durations. The other factors include the quality of the sensory feedback signals from the eyes, the vestibular system, the distal stance limb, the somatosensory system, as well as a successful central nervous system control strategy to coordinate muscle activity (e.g. Ting et. al [17]). One can ask, then, how could the participants, especially the younger adults, balance for four minutes when both measures of hip abduction endurance suggest they should have had a UST less than half of that? We believe this can be explained by a significant contribution to the hip abductor moment from mechanisms other than the active hip abductor muscles themselves. In the next paragraph we explore some of the candidates for reducing the active hip abduction muscle moment required for OLB.

Flamingos rely on passive tissue mechanisms to stand and sleep on one leg [18]. Is it possible that in OLB humans offload their active abductor muscles by stretching passive tissues about the hip to create an added abduction moment? This question was indirectly addressed in the older orthopedic literature when the hip abduction moment required to maintain OLB was calculated as a way of estimating the daily loads and stresses on the femoral head and acetabulum in order to design the first hip endoprostheses with adequate strength. For example, in 1947 Inman estimated the hip abduction moment demand during OLB when standing with a level pelvis and a straight trunk was equal to the weight force of the person times half the distance between the left and right hip joint centers [19]. In 1970, Charnley and McLeish meticulously verified Inman’s calculations experimentally with three subjects, but interestingly showed that the hip abduction moment demand greatly depended on the ‘pelvis attitudes’ [20]. By pelvis attitude they meant the inclination of the pelvis with respect to the horizontal during OLB. However, they never quantified the relationship between the pelvic attitude and the required hip abduction moment. Neither did they express the hip abduction moment required during OLB as a proportion of the maximum voluntary hip abduction isometric strength. So the percentage of hip abduction strength that is required for OLB remained unknown.

### 1) Could there be a systematic bias in our measurements of the parameters that led to an overestimation of the intensity of hip abduction moment?

We do not believe so. In S2 Text we compared our measurements of maximum voluntary hip abduction strength (MVS) and the mass of the body balancing over the stance leg as a percent of total body weight (m_2_) with those from the literature and showed no systematic bias.. We also quantified the errors that we could have had in determining the moment arm of m_2_ and considered all the error to act as a bias contributing to an exaggerated estimation of the intensity. We then showed that even with the most conservative (and unrealistic) way of accounting for these hypothetical biases, the mean calculated intensity for the young men and women would decrease from 53 and 55% to 38 and 45%, respectively. Even at 38% of “allegedly unbiased” intensity the endurance time model for the knee and trunk would anticipate a maximum UST no longer than 120 s and 204 s, respectively. Given that the participants who were halted at the four minutes could still have continued standing on one leg, this clearly shows that all the discrepancy between the anticipated endurance time and UST cannot be attributed to errors in estimation of the intensity of the hip abduction moment demand.

### 2) Could large variations in intensity of hip abduction moment demand during the OLB trial allow time for recovery from the effects of hip abductor muscle fatigue?

Because the mean slope of the trend line passing through the time-series for the intensity of hip abduction moment demand during the OLB trial (**Table 2**) was not significantly different from zero for any of our groups, we do not believe there was enough time for abductor recovery. In addition, the average root mean square of error (RMSE) between the trend line and the time-series for intensity was less than 4%. Therefore, we believe these variations did not allow recovery from fatiguing of the hip abductor muscles.

### 3) Could the lateral bending of the trunk away from the stance hip have relieved the demand on the hip abductor muscles during OLB?

In our earlier study [10] we calculated that lateral flexion of the trunk through half its maximum range of motion could reduce the intensity of the hip abduction moment demand during OLB by 37%. Should the participants have maintained their trunk lateral flexion, that effect would not have been captured by the inverse dynamic algorithm calculating the hip abduction moment because the double inverted pendulum model assumed, for simplicity, that the upper body and the non-stance leg were a single rigid body (M_2_). To account for this source of error, we calculated the time-series of the parameters describing the rigid body M_2_ and observed that they do not have large sudden changes, then chose their trimmed mean value as the parameters in the inverse dynamics equations. We also examined the time-series data for the lateral bending of the trunk for each subject (Table 2 in S2 Text). We noticed that on average our participants did slightly laterally flex their trunk as the OLB trial proceeded (95% Mean Confidence Interval of 1.5 and 3.5 deg), the effect of which would be negligible in the calculation of intensity.

### 4) Could out of plane rotation of the upper body towards the stance limb increase the moment arms of the hip abductor muscles and therefore reduce the hip abduction moment demand of OLB?

There are published modeling data that show that internal and external rotation of the hip changes the moment arms of the hip abductor muscles [21]. So if a participant keeps adjusting about the vertical axis, they could technically be changing their hip abductor muscles’ moment arms and perhaps relieve some of the fatigue. But this possibility can be eliminated because the participants were instructed and monitored to face forwards at all times during their OLB and not twist around the vertical axis. In addition, we used the least square method to pass a vertical plane through the two hip joints centers and the stance leg knee and ankle. We then examined the time-series of the angle between the defined plane and the lab’s frontal plane as a proxy for the hip internal and external rotation angle. We noticed that the slope of the trend line passing through the time-series was not significant and its RMSE were under 2 degrees. Therefore, we can conclude that this could not be a substantial contributor to relieving the abduction moment intensity for our participants during the OLB trial.

### 5) Could the hip abductor muscle fiber composition be more fatigue resistant than the adjacent knee and trunk muscles?

It is possible, but unlikely, that the hip abductor muscles have a higher type I muscle fiber composition than the knee and trunk muscles that we used to estimate endurance times [22,23].

However, our measurements of each participant’s own endurance time at 50% of their measured maximum hip abduction strength points to the same conclusion that their UST was significantly longer than their measured hip abduction muscle endurance time (**Fig 3A)**.

#### What is the least amount of passive tissue contribution as a percentage of the hip abduction moment demand during OLB?

To stand on one leg for more than four minutes, one can calculate that a young person should not be using more than 26% of their hip abduction MVS (based on published endurance time – intensity models of the trunk muscle [14]). In our experiment we calculated the intensity of the hip abduction demand during OLB to be 53% and 55% MVS in young men and women, respectively. One can then estimate that at least 51% (=1 – 26% / 53%) and 53% of the hip abduction demand should be supported by passive tissues stretched by muscles other than abductors. Inman estimated the hip abductor muscle moment during OLB by comparing the EMG activity in abductor muscles while standing on one leg vs during maximal abduction exertion [19]. In the S1 Table, we reused his estimates of the abductor muscles’ torque and OLB abduction moment and calculated that on average 47% and 44% of the abduction moment in OLB (men and women respectively) has to come from a source other than the abductor muscles of the stance hip.

Limitations of the present study were that we did not use electromyography to monitor the state of fatigue of the hip abductor muscles. However, this would have required wire electrodes to reach gluteus medius and minimus, and even if we had done so, the small sampling volumes might not have been representative of the whole muscle activity (for example, Basmajian et. al [24]). While the method for measuring hip abduction endurance in the supine and gravity free plane had the advantage that it did remove any gravitational bias on the hip abductor MVS value, it had the drawback that one had to support the weight of the leg on a frictionless swing arm, and also provide rigid reaction force supports for the pelvis and torso in the horizontal plane. In retrospect it might have been better to measure abduction strength while standing on one leg with a strap and series force transducer around both ankles and one hand supported by a handrail. A further limitation was that we had to utilize muscle endurance time models from the knee and trunk because no data were available for the hip. Given that trunk was the second most fatigue resistive model [14], we may have actually overestimated hip abduction endurance time making it more, not less, difficult for us to reject the main hypothesis.

## 6. Conclusions

The hip abduction moment demand during OLB is strenuous, exceeding more than 50% of the maximum hip abduction strength available, whether in healthy young or older males or females. But even for an essentially level pelvis, we conclude that nearly half of the hip abduction moment developed arises from the stretch of passive tissues creating an abduction moment about the hip. Exactly which tissues those are and the mechanism by which they are stretched remains an open question.

## Data Availability

Aggregated relevant data are available in the Supporting Information files in the form of two excel sheets.

## 7. Acknowledgments

## 9. Supporting information captions

**S1 Text. Detailed methods for the one-legged balance experiment**

**S2 Text. Quantifying the sensitivity of the calculated mean intensity of the hip abduction moment demand to potential biases in the calculations**

**S1 Table. Contribution of tissues other than hip abductor muscles for supporting hip abduction moment demand during one-legged balance**. Calculated by using Inman’s (1947) estimates of the abductor muscles contribution to supporting the abduction moment demand during one-legged stance based on myography measurements of the muscles.

**S2 Table. OLB Subject Parameters**

**S3 Table. OLB MoCap Aggregates**

